# Medical students in distress: a mixed methods approach to understanding the impact of debt on well-being

**DOI:** 10.1101/2024.02.05.24302356

**Authors:** Adrienne Yang, Simone Langness, Lara Chehab, Nikhil Rajapuram, Li Zhang, Amanda Sammann

**Affiliations:** Department of Surgery, University of California, San Francisco, CA; Department of Trauma Surgery, Sharp HealthCare, San Diego, CA; Department of Pediatrics, Stanford University, Stanford, CA; Department of Epidemiology and Biostatistics, University of California, San Francisco, CA

## Abstract

**Background:** Nearly three in four U.S. medical students graduate with debt in six-figure dollar amounts which impairs students emotionally and academically and impacts their career choices and lives long after graduation. Schools have yet to develop systems-level solutions to address the impact of debt on students’ well-being. The objectives of this study were to identify students at highest risk for debt-related stress, define the impact on medical students’ well-being, and to identify opportunities for intervention.

**Methods:** This was a mixed methods, cross-sectional study that used quantitative survey analysis and human-centered design (HCD). We performed a secondary analysis on a national multi-institutional survey on medical student wellbeing, including univariate and multivariate logistic regression, a comparison of logistic regression models with interaction terms, and analysis of free text responses. We also conducted semi-structured interviews with a sample of medical student respondents and non-student stakeholders to develop insights and design opportunities.

**Results:** Independent risk factors for high debt-related stress included pre-clinical year (OR 1.75), underrepresented minority (OR 1.40), debt $20-100K (OR 4.85), debt >$100K (OR 13.22), private school (OR 1.45), West Coast region (OR 1.57), and consideration of a leave of absence for wellbeing (OR 1.48). Mental health resource utilization (p= 0.968) and counselors (p= 0.640) were not protective factors against debt-related stress. HCD analysis produced 6 key insights providing additional context to the quantitative findings, and associated opportunities for intervention.

**Conclusions:** We used an innovative combination of quantitative survey analysis and in-depth HCD exploration to develop a multi-dimensional understanding of debt-related stress among medical students. This approach allowed us to identify significant risk factors impacting medical students experiencing debt-related stress, while providing context through stakeholder voices to identify opportunities for system-level solutions.

## Introduction

Over the past few decades, it has become increasingly costly for aspiring physicians to attend medical school and pursue a career in medicine. Most recent data shows that 73% of medical students graduate with debt often amounting to six-figures^1^ – an amount that is steadily increasing every year.^2^ In 2020, the median cost of a four-year medical education in the United States (U.S.) was $250,222 for public and $330,180 for private school students^1^ – a price that excludes collateral costs such as living, food, and lifestyle expenses. To meet these varied costs, students typically rely on financial support from their families, personal means, scholarships, or loans. Students are thereby graduating with more debt than ever before and staying indebted for longer, taking 10 to 20 years to repay their student loans regardless of specialty choice or residency length.^1^

Unsurprisingly, higher debt burden has been negatively correlated with generalized severe distress among medical students,^3,4^ in turn jeopardizing their academic performance and potentially impacting their career choices.^5^ Studies have found that medical students with higher debt relative to their peers were more likely to choose a specialty with a higher average annual income,^5^ less likely to plan to practice in underserved locations, and less likely to choose primary care specialties.^4^ However, a survey of 2019 graduating medical students from 142 medical schools found that, when asked to rank factors that influenced their specialty choice, students ranked economic factors, including debt and income, at the bottom of the list. With this inconsistency in the literature, authors Youngclaus and Fresne declare that further studies and analysis are required to better understand this important relationship.^1^

Unfortunately, debt and its negative effects disproportionately impact underrepresented minority (URM) students, including African Americans, Hispanic Americans, American Indian, Native Hawaiian, and Alaska Native,^6^ who generally have more debt than students who are White or Asian American.^1^ In 2019, among medical school graduates who identified as Black, 91% reported having education debt, in comparison to the 73% reported by all graduates.^1^ Additionally, Black medical school graduates experience a higher median education debt amount relative to other groups of students, with a median debt of $230,000.^1^ This inequitable distribution of debt disproportionately places financial-related stress on URM students,^7^ discouraging students from pursuing a medical education.^8^ These deterring factors can lead to a physician workforce that lacks diversity and compromises health equity outcomes.^9^

Limited literature exists to identify the impact of moderating variables on the relationship between debt and debt-related stress. Financial knowledge is found to be a strong predictor of self-efficacy and confidence in students’ financial management, leading to financial optimism and potentially alleviating debt stress.^10–12^ Numerous studies list mindfulness practices, exercise, and connecting with loved ones as activities that promote well-being and reduce generalized stress among students.^13–15^ However, to date, no studies have examined whether these types of stress-reducing activities, by alleviating generalized stress, reduce debt-related stress. Studies have not examined whether resources such as physician role models may act as a protective factor against debt-related stress.

Despite the growing recognition that debt burdens medical students emotionally and academically, we have yet to develop systemic solutions that target students’ unmet needs in this space. We performed the first multi-institutional national study on generalized stress among medical students, and found that debt burden was one of several risk factors for generalized stress among medical students.^3^ The goal of this study is to build upon our findings by using a mixed methods approach combining rigorous survey analysis and human-centered design to develop an in-depth understanding of the impact that education debt has on medical students’ emotional and academic well-being and to identify opportunities for intervention.

## Methods

We conducted a mixed methods, cross-sectional study that explored the impact of debt-related stress on US medical students’ well-being and professional development. This study was conducted at the University of California, San Francisco (UCSF). All activities were approved by the UCSF institutional review board. We performed a secondary analysis of the quantitative and qualitative results of the Medical Student Wellbeing Survey (MSWS), a national multi-institutional survey on medical student wellbeing administered between 2019-2020, to determine risk factors and moderating variables of debt-related stress. To further explore these variables, we used human-centered design (HCD), an approach to problem-solving that places users at the center of the research process in order to determine key pain points and unmet needs, and co-design solutions tailored to their unique context.^16^ In this study, we performed in-depth, semi-structured interviews with a purposefully sampled cohort of medical students and a convenience sample of non-student stakeholders to determine key insights representing students’ unmet needs, and identified opportunities to ameliorate the impact of debt-related stress on medical students.

### Quantitative Data: The Medical Student Wellbeing Survey

The MSWS is a survey to assess medical student wellbeing that was administered from September 2019 to February 2020 to medical students actively enrolled in accredited US or Caribbean medical schools.^3^ Respondents of the MSWS represent a national cohort of >3,000 medical students from >100 unique medical school programs. The MSWS utilizes a combination of validated survey questions, such as the Medical Student Wellbeing Index (MS-WBI), and questions based on foundations established from previously validated wellbeing survey methods.^3^ Questions generally focused on student demographics, sources of stress during medical school, specialty consideration, and frequency in activities that promote wellbeing.

Some questions ask students to rate physical, emotional, and social domains of wellbeing using a five-point Likert scale. Questions of interest from the MSWS included debt-related stress, generalized stress, intended specialty choice, and utilization of well-being resources and counselors. An additional variable investigated was average school tuition, which was determined by a review of publicly available data for each student’s listed medical school.^17^

#### Stress: Debt-Related and Generalized Stress

Debt stress was assessed by the question, “How does financial debt affect your stress level?” Students responded using a five-point Likert scale from –2 to 2: significant increase in stress (−2), mild increase (−1), no change (0), mild decrease (1), or significant decrease (2).

Responses for this question were evaluated as a binary index of ‘high debt stress,’ defined as a response of –2, versus ‘low debt stress,’ defined as a response of –1 or 0. In addition, generalized stress from the MSWS was assessed by questions from the embedded MS-WBI, which produced a score. Previous studies have shown that the score can be used to create a binary index of distress: a score ≥ has been associated with severe distress, and a score <4 has been associated with no severe distress.^18^

#### Intended Specialty

We categorized students’ responses to intended specialty choice by competitiveness, using the 2018 National Resident Match Program data.^19^ ‘High’ and ‘low’ competitiveness were defined as an average United States Medical Licensing Examination (USMLE) Step 1 score of >240 or ≤230, respectively, or if >18% or <4% of applicants were unmatched, respectively. ‘Moderate’ competition was defined as any specialty not meeting criteria for either ‘high’ or ‘low’ competitiveness.

#### Resource Utilization

The MSWS assessed utilization of well-being resources by the question, “At your institution, which of the following well-being resources have you utilized? (Select all that apply)” Students responded by selecting each of the resource(s) they used: Mental Health and Counseling Services, Peer Mentorship, Self-Care Education, Mindfulness/Meditation Classes, Community Building Events, and Other. The number of choices that the student selected was calculated, allowing for placement into a category depending on the amount of resource utilization: 0-20%, 20-40%, 40-60%, 60-80%, 80-100%. Responses for this question were evaluated as a binary index of ‘high resource utilization,’ defined as a response of 80-100% resource utilization, versus ‘low resource utilization,’ defined as a response of <80% resource utilization. Additionally, use of a counselor for mental health support was assessed by the question, “Which of the following activities do you use to cope with difficult situations (or a difficult day on clinical rotation)? (Select all that apply)” Students responded by selecting the activities that they use from a list, which included: Listen to Music, Mindfulness Practice, Meet with a Counselor, Exercise, etc. Responses for this question were evaluated as a binary index of ‘Meeting with a Counselor,’ defined by selection of that option, versus ‘Not Meeting with a Counselor,’ defined as not selecting that option.

### Quantitative Data Analysis

We performed a secondary analysis of quantitative data from the MSWS to calculate frequencies and odds ratios for the five quantitative variables described above (debt-related stress, generalized stress, intended specialty, resource utilization, and school tuition). Univariate analysis and multivariate logistic regression were performed among students in the high debt stress (−2) and low debt stress (0 or –1) for select variables, such as clinical phase, URM, debt burden, specialty competitiveness, and average school tuition among others to identify risk factors for high debt stress. To determine if ‘high resource utilization’ or ‘meeting with a counselor’ were moderating variables on the relationship between debt burden and debt stress, we applied the logistic regression with the interaction terms of ‘debt’ and ‘resource utilization’ (high vs. low). Then, we performed a similar analysis but replaced the interaction term with ‘debt’ and ‘meeting with a counselor’ (yes vs. no). We also performed Chi-squared tests to determine the degree to which severe distress increases as debt burden increases, if specialty competitiveness varied by debt stress, and if the proportion of students who identified as URM, in comparison to non-URM, differed by debt level. All statistical tests were two-sided and p <0.05 was considered significant. Statistical analyses were performed using SAS version 9.4 and R version 4.0.5.

### Qualitative Data: Interviews and MSWS Free Text Responses

#### Free-Text Entries

At the conclusion of the 2019-2020 MSWS, respondents had unlimited text space to provide comments to two prompts. The first prompt read, “What well-being resource(s), if offered at your school, do you feel would be most useful?” The second prompt read “If you have any further comments to share, please write them below.” Answers to either prompt that pertained to debt, cost of medical school, or finances were extracted for the purpose of this study and analyzed with the other qualitative data subsequently described.

#### Interview Selection & Purposive Sampling

Interview participants were identified from a repository of respondents to the MSWS who had attached their email address and expressed willingness at the time of the survey to be contacted for an interview.^3^ Our recruitment period was between April 19, 2021 to July 2, 2021. The recruitment process involved sending invitations to all of the email addresses in the list to participate in a 45-minute interview on the topic of student debt and wellbeing. The invitation included a brief screening questionnaire asking students to report updates to questions that were previously asked in the MSWS (i.e.: clinical training year, marital status, dependents). Additional novel questions included primary financial support system, estimate of financial support systems’ household income in the last year, estimate of educational financial debt at conclusion of medical school, student’s plan for paying off debt, and degree of stress (using a Likert scale from 0-10) over current and future education debt.

Purposeful sampling of medical student stakeholders for interviews allowed us to maximize heterogeneity. We utilized the students’ responses to the brief screening questionnaire with their corresponding responses to demographic questions from the MSWS to select interviewees that varied by gender, race, presence of severe distress, type of medical school (public vs. private), region of school, and tuition level of school. The sampling ensured a diverse representation, in accordance with HCD methodology.^20^ Brief descriptions of participant experiences are listed in **Table 2** (“Interviewee Descriptors”). Students who were selected for interviews were sent a confirmation email to participate. In addition, to include representation from the entire ecosystem, we interviewed a financial aid counselor at a medical school and a pre-medical student, chosen through convenience sampling. We directly contacted those two individuals for interviews.

#### Semi-Structured Interviews

All interviews were conducted between April 2021 and July 2021 over Zoom, a collaborative, cloud-based videoconferencing service offering secure audio recordings of sessions. A single researcher conducted interviews over an average of 45 minutes. Informed consent was obtained verbally from participants prior to interviews; interviews and their recordings only proceeded following verbal consent. The interview guide (S1 File) included open-ended questions about students’ experience of debt-related stress and their reflections on its consequences. The audio recordings were transcribed using Otter.ai, a secure online transcription service that converts audio files to searchable text files. Interview responses were redacted to preserve anonymity of respondent identity.

#### Qualitative Data Analysis

Interview data was analyzed using a general inductive approach to thematic analysis. Specifically, two researchers (SL and AY) independently inductively analyzed transcripts from the first three semi-structured interviews to come up with themes relating to the experiences and consequences of debt-related stress. They reconciled discrepancies in themes through discussion to create the codebook (S2 File), which included 18 themes. SL and AY independently coded each subsequent interview transcript as well as the free text responses from the survey, meeting to reach a consensus on representative quotes for applicable themes.

The two researchers met with the core research team to extrapolate ‘insight statements’ from the themes that emerged from the thirteen interviews, deducing unique human perspectives, motivations, or tensions from the thematic data following the HCD methodology.^21^ These insight statements articulate key challenges experienced by stakeholders, and provide the basis for design opportunities that lead to collaborative co-design processes for solution development. In HCD, design opportunities are statements derived directly from the insight and are framed in a way that suggest that multiple solutions are possible. These design opportunities are used during brainstorming sessions, where creativity and innovation are encouraged in the co-design process.^22^

## Results

### MSWS Respondents and Quantitative Analysis

A total of 3,162 students responded to the MSWS and their sociodemographic characteristics have been described previously.^3^ A total of 2,771 respondents (87.6%) provided responses for our study’s variables of interest, including a response for ‘high debt stress’ (–2) or ‘low debt stress’ (–1 or 0). **Table 1** lists the distribution of debt-related stress across different variables for all respondents.

**Table 1.**
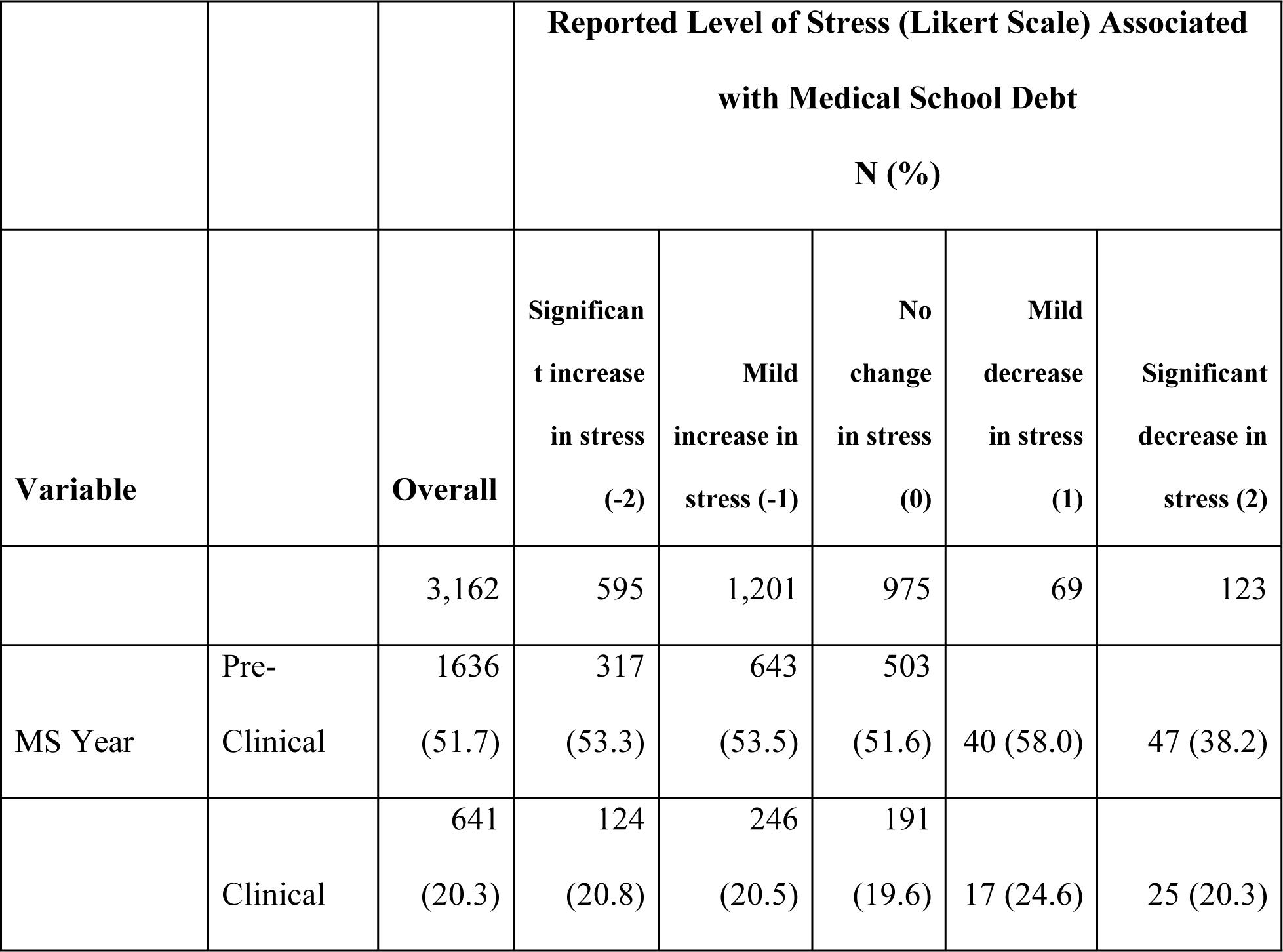
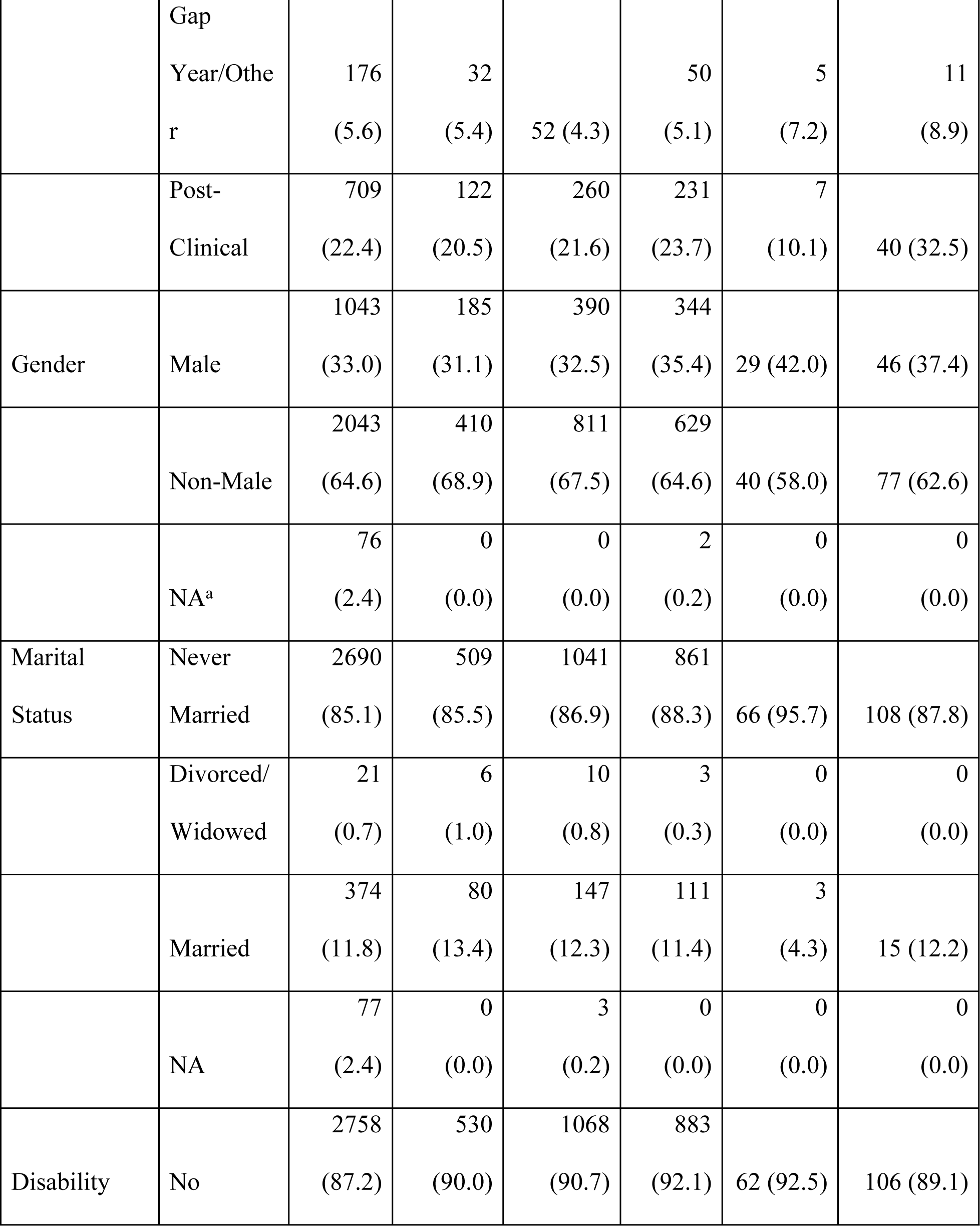
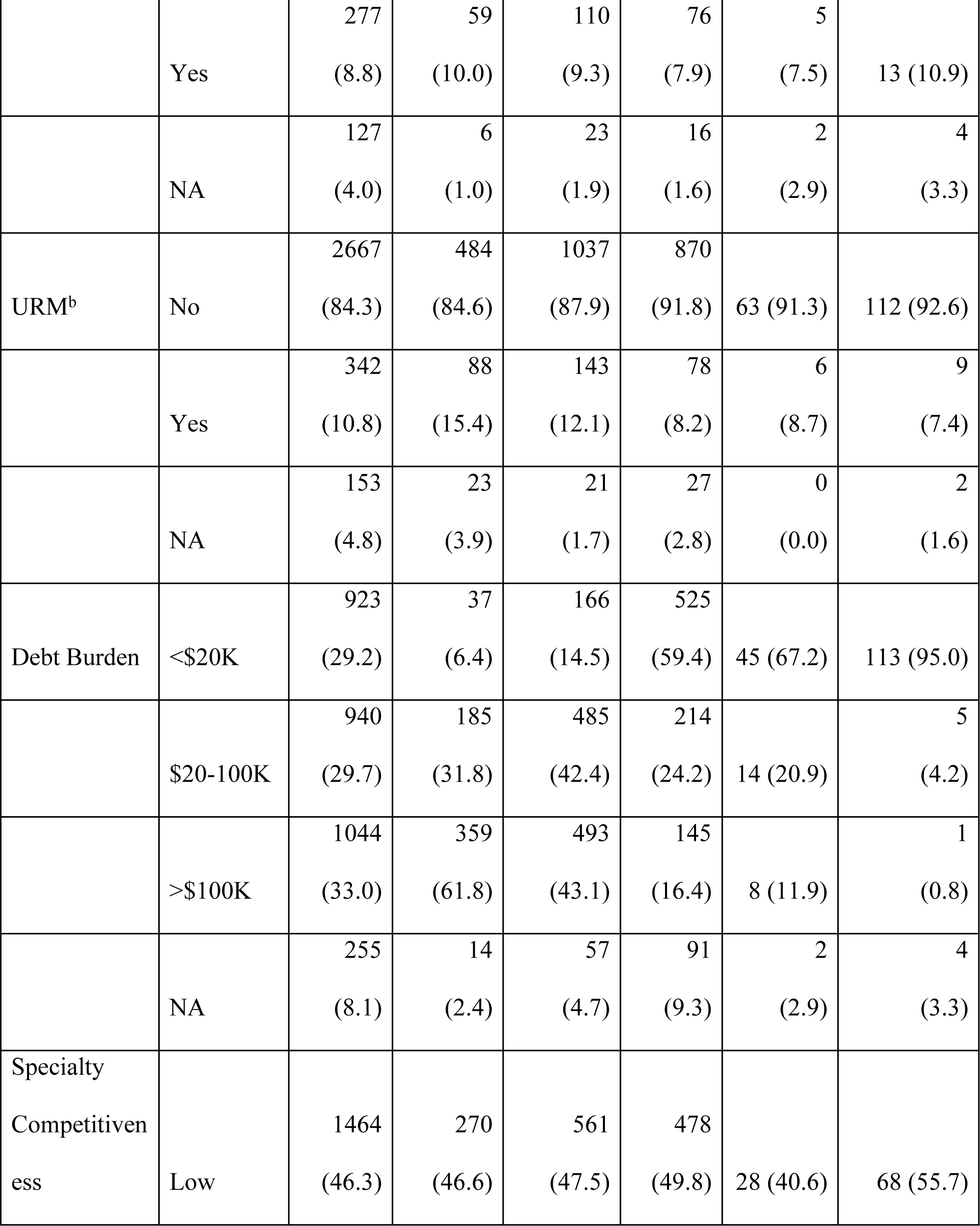
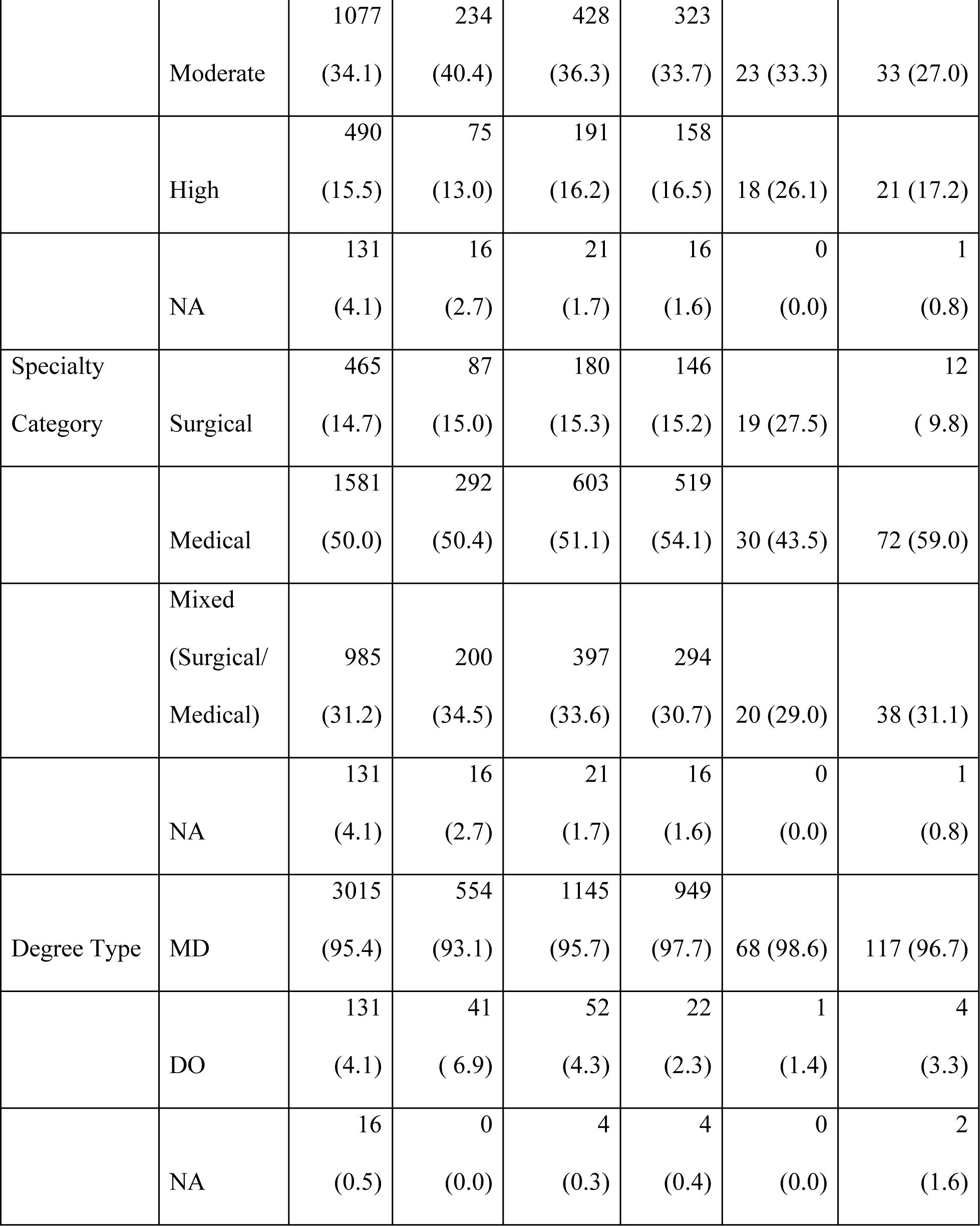
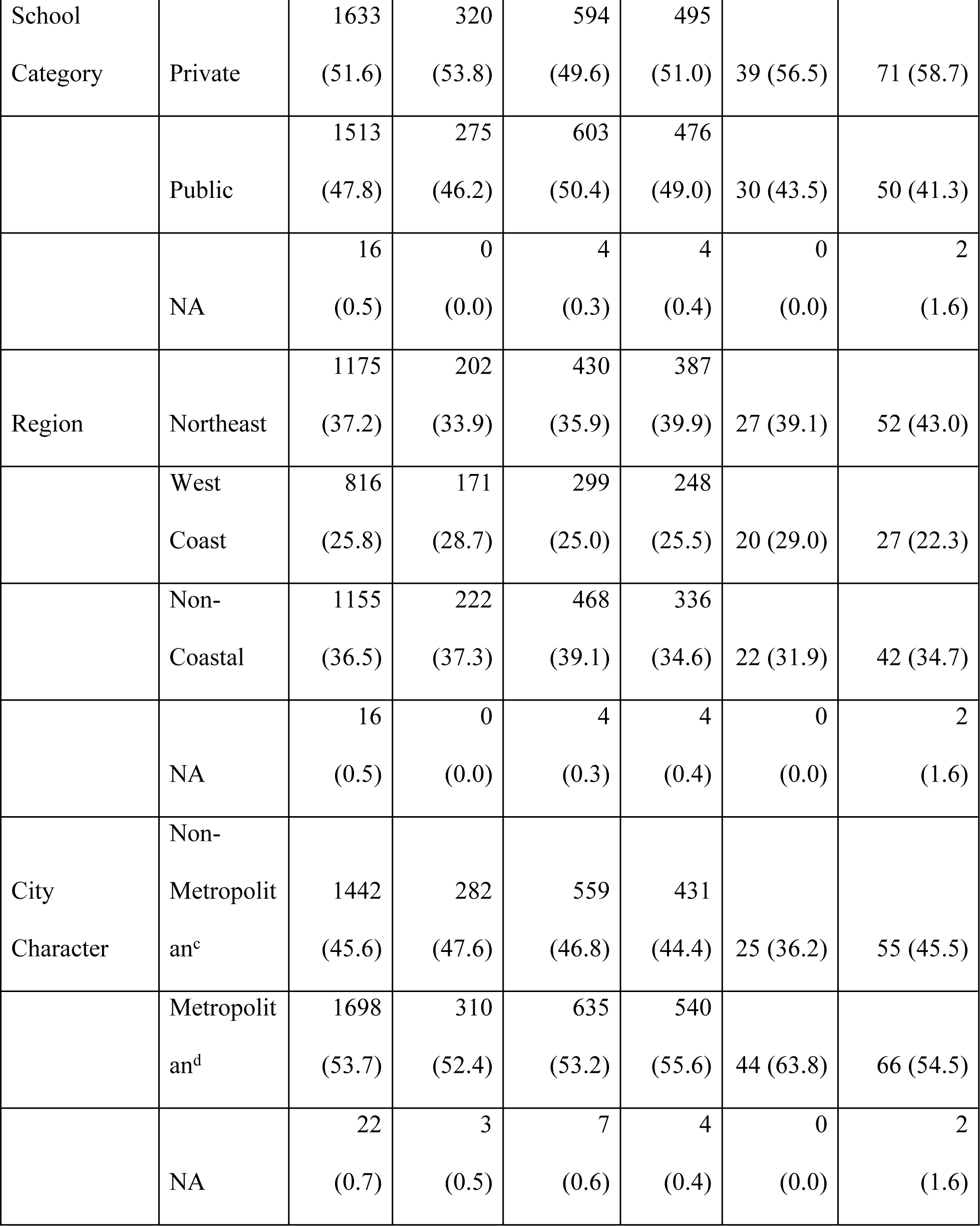
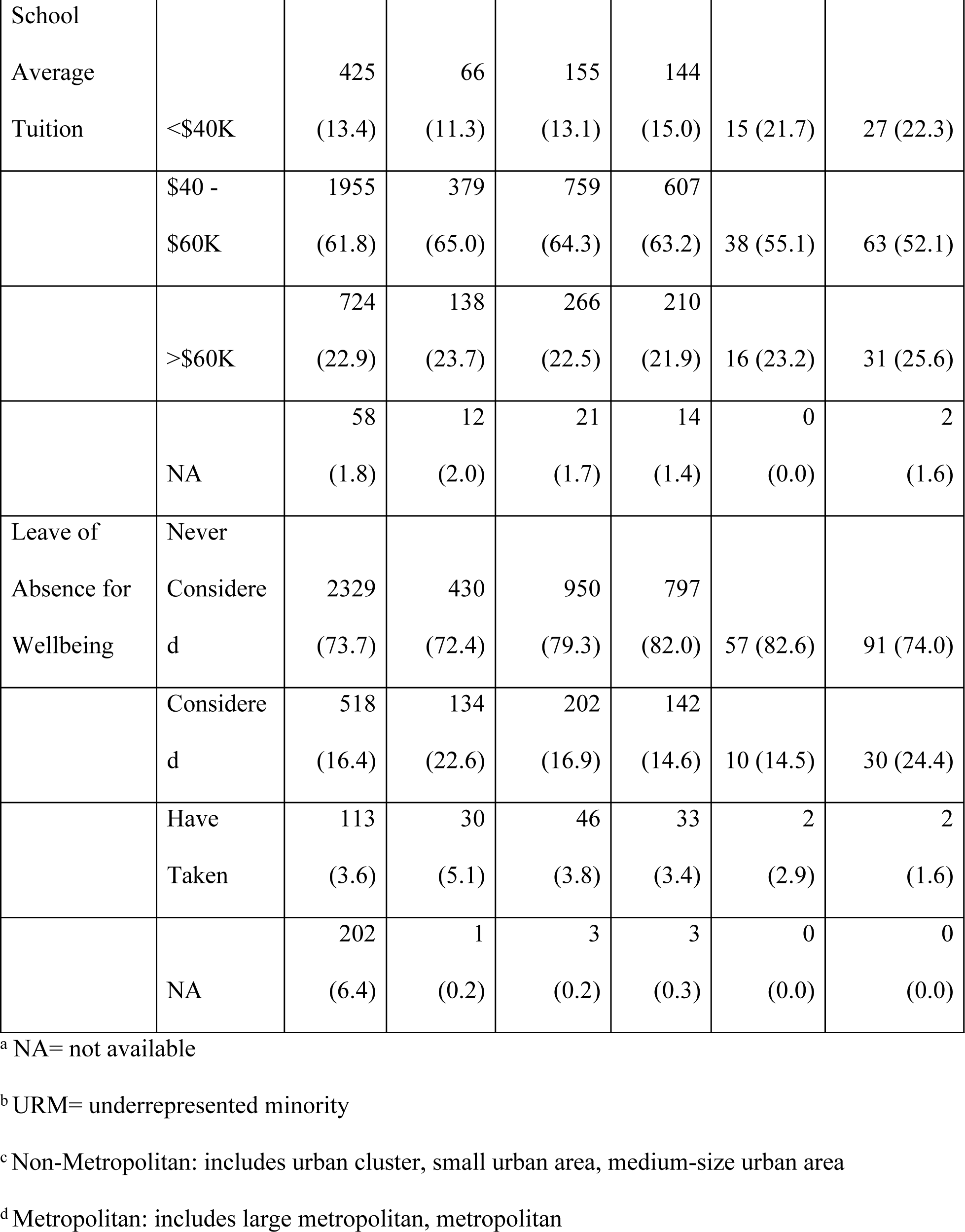
Summary statistics of debt stress level for different variables (N=3,162)

**Table 2.** Interviewee Descriptors.

|  |  | Summary of Interviewees |
| --- | --- | --- |
| Student Stakeholders | 1 | 2nd-year male student with high debt stress, who is married and pursuing a highly competitive specialty |
|  | 2 | 2nd-year female student with low debt stress, who comes from a low-income background with receives financial support |
|  | 3 | Gap year (PhD years of MD/PhD program) female student with low debt stress, who comes from a wealthy background and is pursuing a lower paying specialty |
|  | 4 | 2nd-year female immigrant student with high debt stress, pursuing a lower paying specialty and a career of advocacy |
|  | 5 | 3rd-year female student with low debt stress, who is married and receives financial support |
|  | 6 | 4th-year female student with low debt stress, who identifies as URM and is graduating debt-free due to financial aid |
|  | 7 | 3rd-year male student with low debt stress, who is enrolled in an MD/PhD program and is receiving financial support from his fiancée |
|  | 8 | 4th-year male student with low debt stress, who took out loans and is pursuing a higher paying specialty |
|  | 9 | 2nd-year female student with low debt stress, who took out loans and is enrolled in a 7-year medical program (including college and medical school) |
|  | 10 | 1st-year female student with low debt stress, who receives scholarships for medical school and whose parent is a physician |
|  | 11 | 1st-year female immigrant student with low debt stress, who identifies as URM and is pursuing a lower paying specialty |
| Non-Student Stakeholders | 12 | Pre-medical female student with high debt stress, who identifies as a "career changer" and is enrolled in a postbaccalaureate program |
|  | 13 | Medical school financial aid counselor |

#### Risk Factors for Debt-Related Stress

Factors that were independently associated with higher debt-related stress included being in pre-clinical year (OR 1.75, 95% CI 1.30-2.36, p<0.001), identifying as URM (OR 1.40, 95% CI 1.03-1.88), p=0.029), having debt $20-100K (OR 4.85, 95% CI 3.32-7.30, p<0.001), debt >100K (OR 13.22, 95% CI 9.05-19.90, p<0.001), attending a private medical school (OR 1.45, 95% CI 1.06-1.98, p=0.019), attending medical school on the West Coast (OR 1.57, 95% CI 1.17-2.13, p=0.003), and having considered taking a leave of absence for wellbeing (OR 1.48, 95% CI 1.13-1.93, p=0.004) (**Table 3, S1 Table).**

**Table 3.** Predictors of high debt stress in medical students using a multivariable logistic regression model (N=2,771)

|  | Univariate Analysis |  | Multivariate Analysis |  |
| --- | --- | --- | --- | --- |
|  | Odds Ratio<br>(95% CI) | P-Value | Odds Ratio<br>(95% CI) | P-Value |
| **Medical School Year (vs. Clinical)** |  |  |  |  |
| Pre-clinical | 0.97 (0.77-1.24) | $p=0.832$ | 1.75 (1.30-2.36) | $p<0.001^*$ |
| Gap Year/Other | 1.11 (0.70-1.71) | $p=0.658$ | 1.09 (0.63-1.86) | $p=0.748$ |
| Post-clinical | 0.88 (0.66-1.16) | $p=0.355$ | 0.85 (0.62-1.17) | $p=0.310$ |
| **Gender (vs. Male)** |  |  |  |  |
| Non-Male | 1.13 (0.93-1.37) | p=0.221 | 1.22 (0.97-1.53) | p=0.085 |
| **Marital Status (vs. Never Married)** |  |  |  |  |
| Married | 1.16 (0.88-1.51) | p=0.284 | 1.07 (0.78-1.45) | p=0.684 |
| Divorced / Widowed | 1.72 (0.60-4.39) | p=0.272 | 1.02 (0.33-2.84) | p=0.973 |
| **Disability (vs. No\*\*)** |  |  |  |  |
| Yes | 1.17 (0.85-1.58) | p=0.324 | 0.95 (0.66-1.34) | p=0.760 |
| **URM (vs. No)** |  |  |  |  |
| Yes | 1.57 (1.20-2.04) | p=0.001<br>* | 1.40 (1.03-1.88) | p=0.029* |
| **Debt Burden (vs. <20K)** |  |  |  |  |
| 20-100K | 4.94 (3.46-7.24) | p<0.001<br>* | 4.85 (3.32-7.30) | p<0.001* |
| >100K | 10.51 (7.47-15.22) | p<0.001<br>* | 13.22 (9.05-19.90) | p<0.001* |
| **Specialty Competitiveness (vs. High)** |  |  |  |  |
| Low | 1.21 (0.92-1.61) | p=0.188 | 1.11 (0.69-1.78) | p=0.669 |
| Moderate | 1.45 (1.09-1.95) | p=0.012<br>* | 1.31 (0.93-1.87) | p=0.131 |
| **Specialty Category (vs. Surgical)** |  |  |  |  |
| Medical | 0.98 (0.75-1.28) | p=0.855 | 1.02 (0.64-1.61) | p=0.940 |
| Mixed | 1.08 (0.82-1.45) | p=0.575 | 1.09 (0.77-1.55) | p=0.621 |
| **Degree Type (vs. MD)** |  |  |  |  |
| DO | 2.09 (1.40-3.09) | p<0.001<br>* | 1.59 (0.91-2.76) | p=0.098 |
| **School Category (vs. Public)** |  |  |  |  |
| Private | 1.15 (0.96-1.38) | p=0.125 | 1.45 (1.06-1.98) | p=0.019* |
| **Region (vs. Non-Coastal)** |  |  |  |  |
| Northeast | 0.90 (0.72-1.11) | p=0.312 | 0.96 (0.72-1.29) | p=0.798 |
| West Coast | 1.13 (0.90-1.42) | p=0.284 | 1.57 (1.17-2.13) | p=0.003* |
| **City Character (vs. Non-Metropolitan)** |  |  |  |  |
| Metropolitan | 0.93 (0.77-1.11) | p=0.409 | 0.92 (0.71-1.18) | p=0.508 |
| **School Average Tuition (vs. <40K)** |  |  |  |  |
| 40-60K | 1.26 (0.95-1.69) | p=0.122 | 0.95 (0.68-1.35) | p=0.774 |
| >60K | 1.31 (0.95-1.83) | p=0.102 | 0.91 (0.60-1.37) | p=0.648 |
| **Leave of Absence for Wellbeing (vs. Never Considered)** |  |  |  |  |
| I have considered taking a leave of absence from school for my personal well-being | 1.58 (1.26-1.98) | p<0.001<br>* | 1.48 (1.13-1.93) | p=0.004* |
| I have taken a leave of absence from school for my personal well-being | 1.54 (0.99-2.35) | p=0.050 | 1.34 (0.78-2.25) | p=0.276 |

\*Statistically significant at p<.05
\*\*Includes: 'no' and 'I'd prefer not to say'

#### Severe Distress by Debt Amount

Levels of generalized severe distress differed across debt burden groups. As debt level increased, the percentage of individuals with “severe” distress increased (p <0.001).

#### Debt and Career Decisions

There were significant differences between the high debt stress versus low debt stress groups and plans to pursue highly vs. moderately vs. minimally competitive specialties (p=.027) **(Fig 1).** A greater percentage of low debt stress students were pursuing a highly competitive specialty or a minimally competitive specialty. A greater percentage of high debt stress students were pursuing a moderately competitive specialty. As shown in **Table 3**, there were no differences in debt-associated stress between students who choose different specialties, such as medical versus surgical versus mixed (medical/surgical).

**Fig 1.**
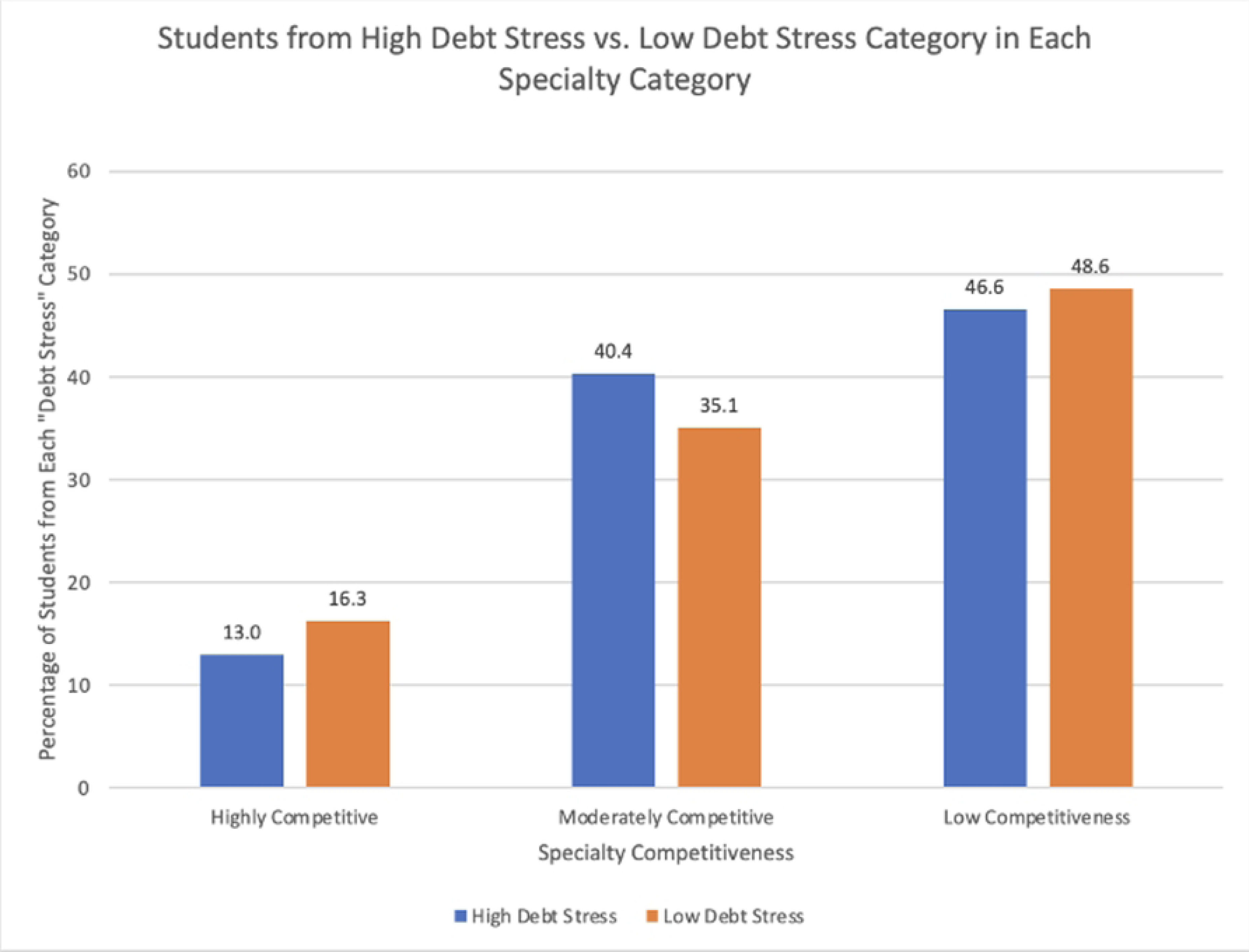
Debt Stress by Specialty Competitiveness.

#### URM Students’ Experience of Debt

URM identity was an independent risk factor for higher debt-related stress **(Table 3).** In addition, debt levels varied between those who identify as URM versus non-URM (p <0.001). Students identifying as URM tended to have higher debt than those who did not. Although the percentage of non-URM students was higher than that of URM students within the lowest debt burden category (<$20k), among all higher debt burden categories, including $20-100K, $100-300K, and >$300K, the percentage of URM students was higher than the percentage of non-URM students.

#### Moderating Factors on the Relationship Between Debt and Debt Stress

Protective factors such as high degree of mental health resource utilization and meeting with a counselor did not reduce the impact of debt burden on debt stress. Among students who reported a high degree of mental health resource utilization, there was no impact on the relationship between debt and debt stress (p= 0.968). Similarly, meeting with a counselor had no impact on the relationship between debt and debt stress (p= 0.640).

### Interview Respondents and Qualitative Analysis

We conducted in-depth, semi-structured interviews with 11 medical students, who are briefly described in **Table 2**. Among the medical student interviewees, there was representation from all regions, including the Northeast (n=3), West Coast (n=5), Midwest (n=2), and South (n=1). Students were also from all clinical phases, including pre-clinical (n=3), clinical (n=4), gap year/other (n=2), and post-clinical (n=2). Most interviewees were female (n=8) and 5 of the interviewees identified as URM. Financial support systems were diverse, including self (n=3), spouse/partner (n=3), and parents/other (n=5). Most interviewees reported low debt stress (n=8), as opposed to high debt stress (n=3). Fifty-five percent of interviewees planned to pursue specialties that pay <$300K (n=6), with some pursuing specialties that pay $300-400K (n=2) and >$400K (n=3).

Among the MSWS free-text responses, to the prompt, “What well-being resource(s), if offered at your school, do you feel would be most useful?” 20 of 118 respondents (16.9%) provided free-text responses that pertained to debt, cost of medical school, or finances. To the prompt “If you have any further comments to share, please write them below” 11 of 342 students (3.2%) provided relevant free-text responses. Analysis of the free-text responses and semi-structured interviews revealed 6 distinct insights **(Table 4)**, with each insight translated into an actionable design opportunity.

**Table 4.**
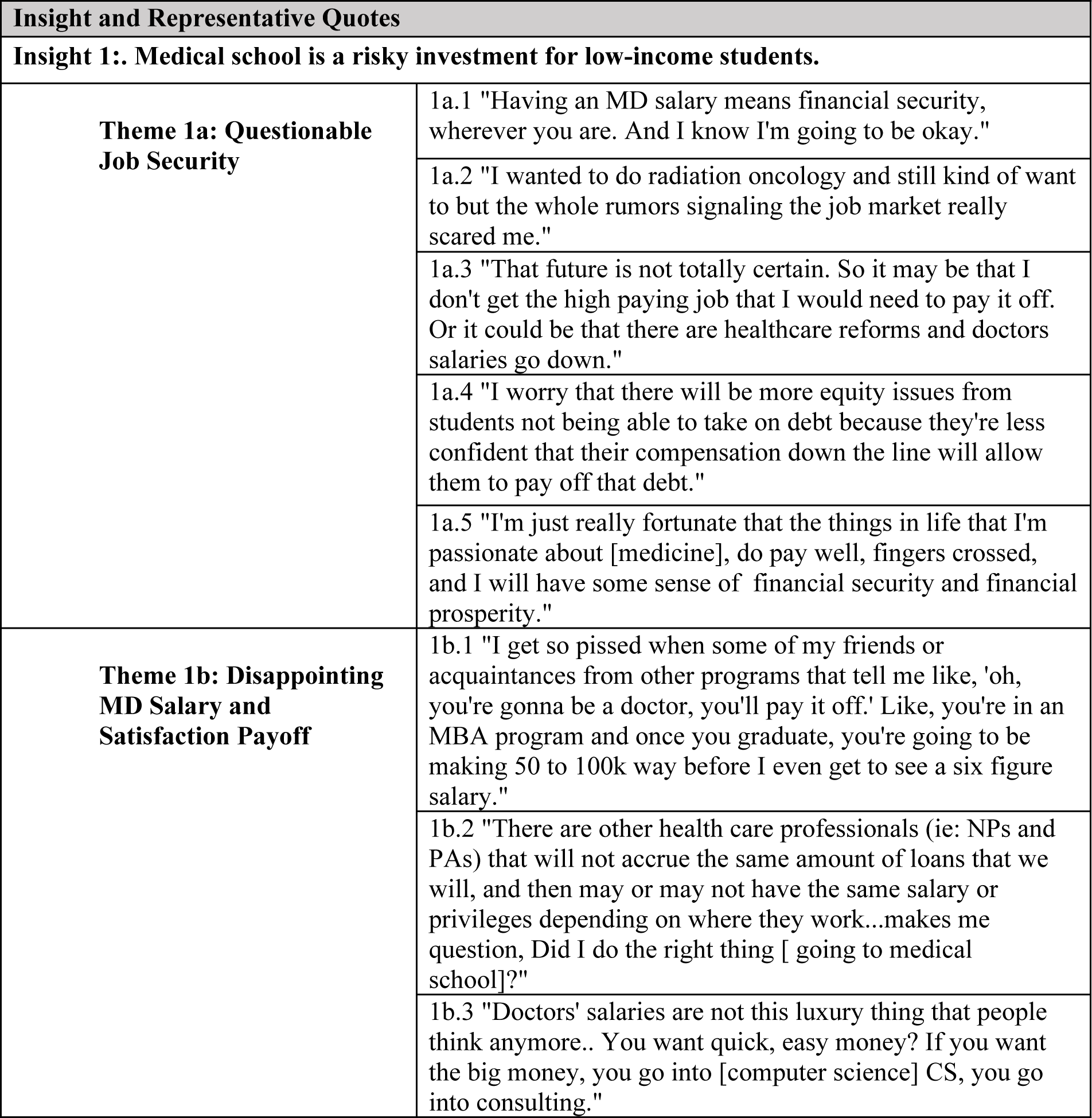
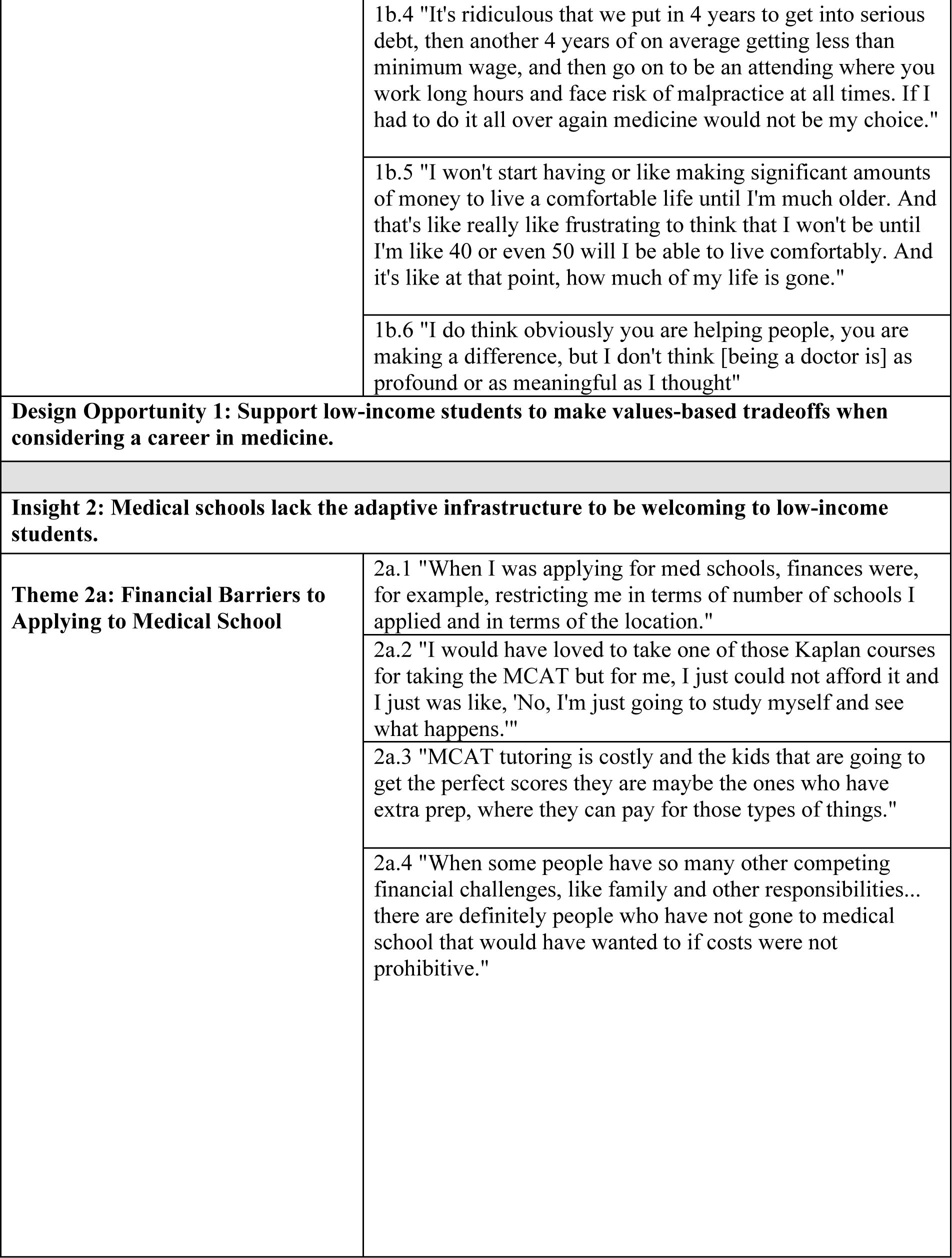
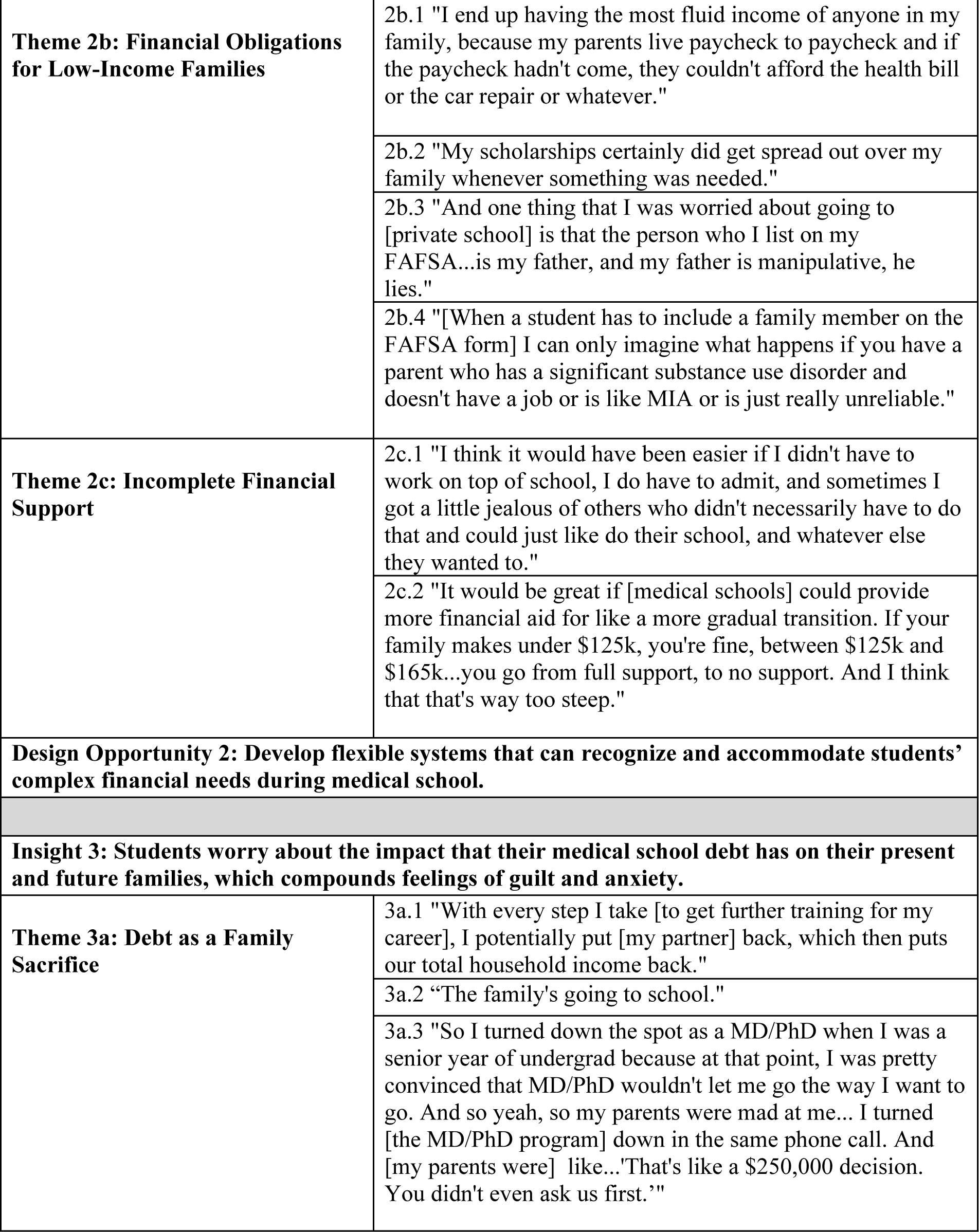
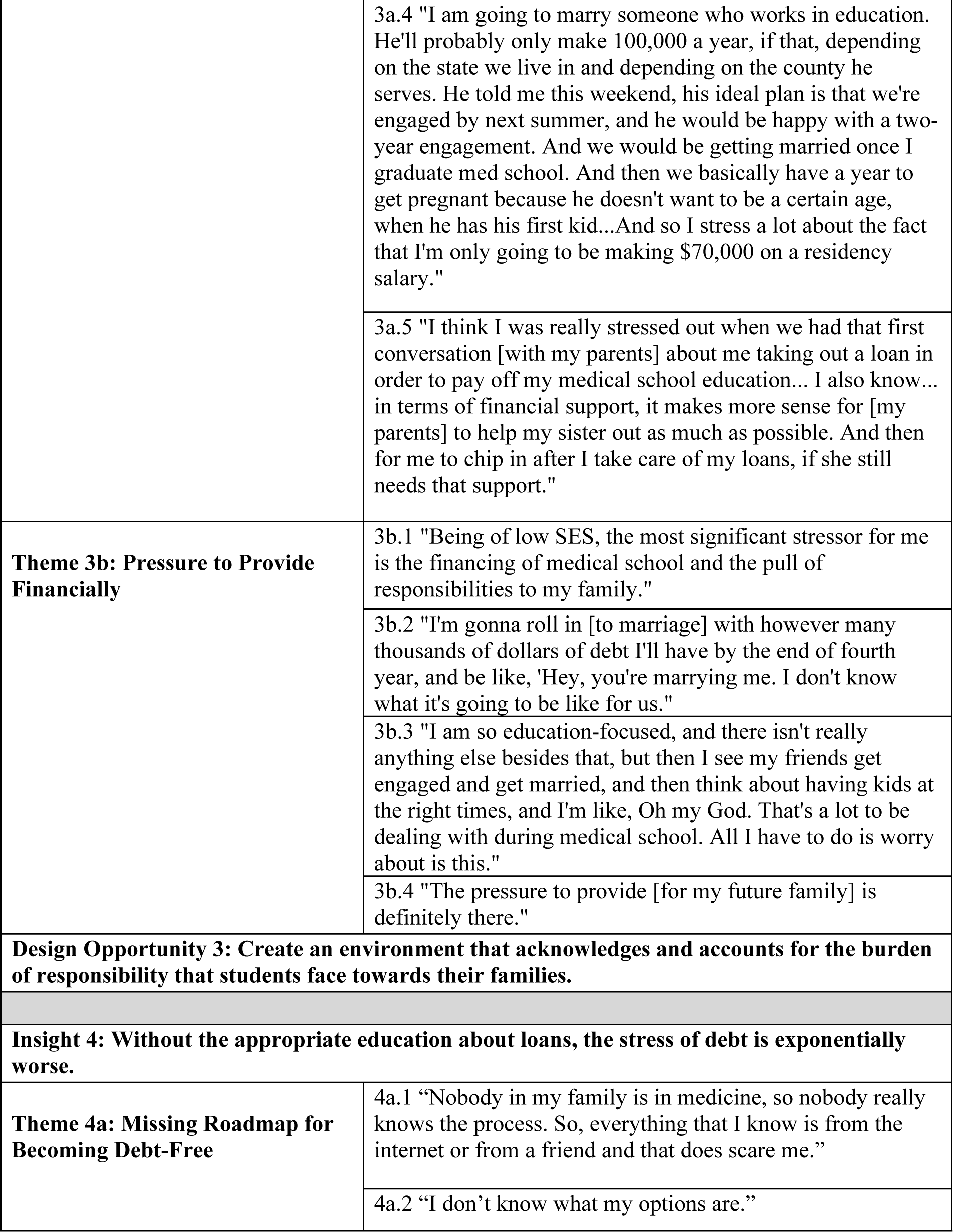
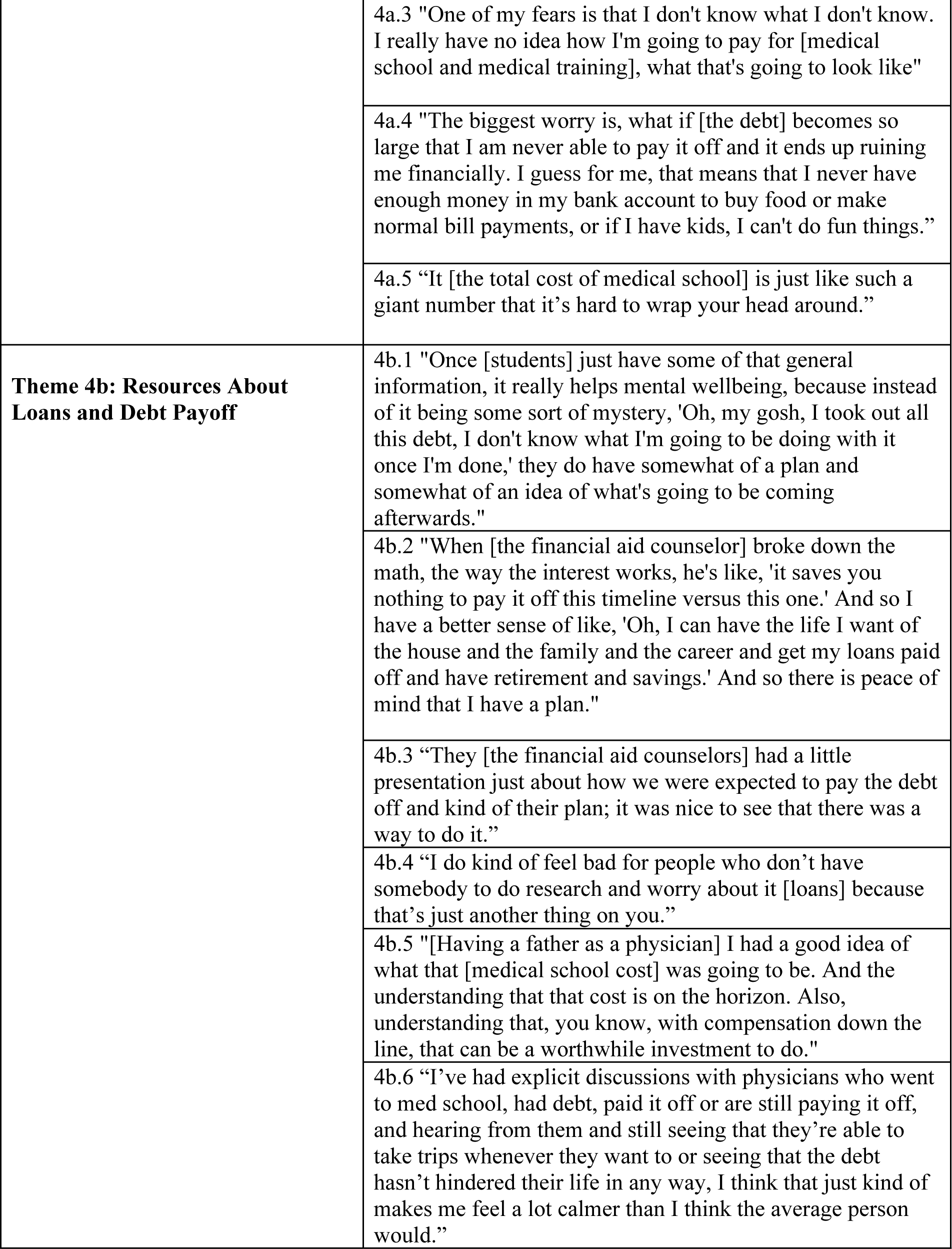
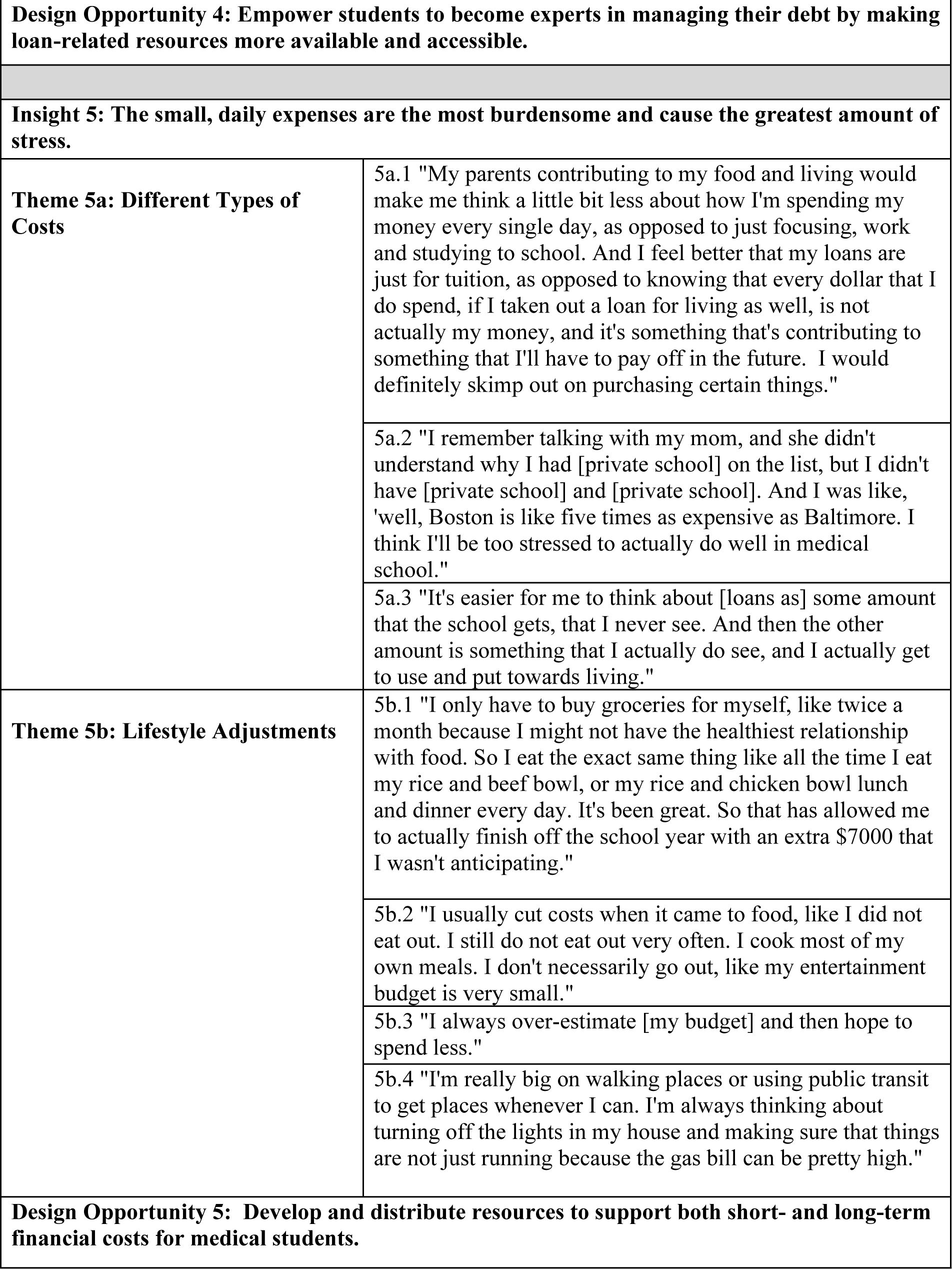
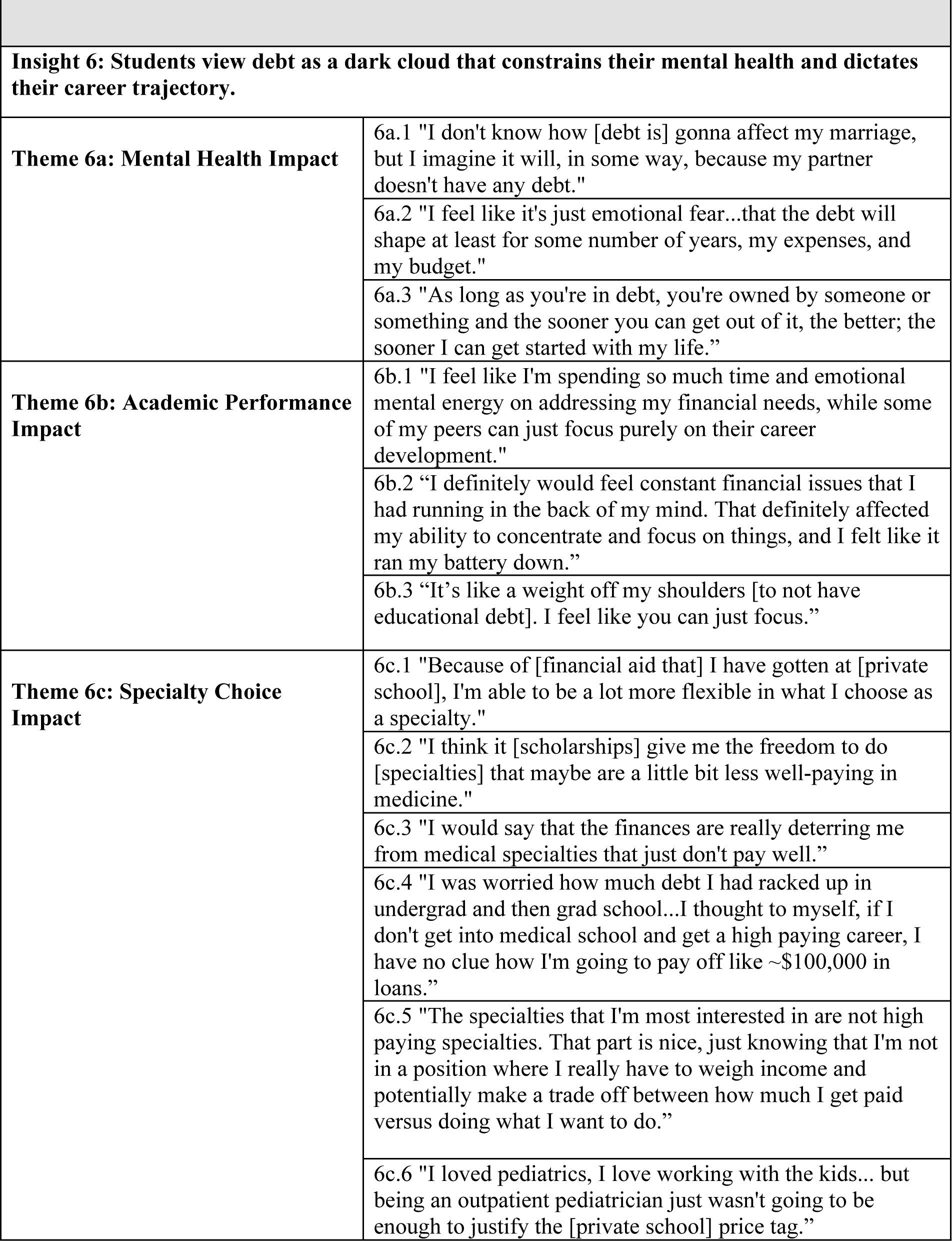
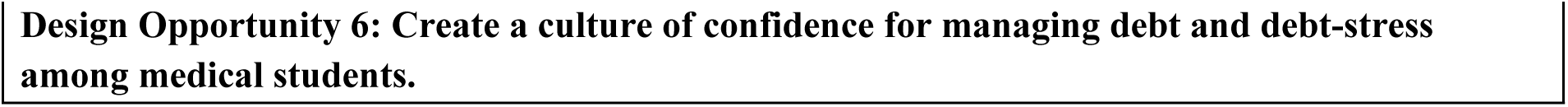
Representative Quotes for Each Insight.

#### Insight 1

Medical school is a risky investment for low-income students.

#### Description

The personal and financial sacrifices required for low-income students to attend medical school and pursue a career in medicine outweigh the benefits of becoming a physician. When considering a career in medicine, students feel discouraged by questionable job security (theme 1a) and reduced financial compensation (theme 1b) – a combination that jeopardizes immediate and long-term job satisfaction. Some students feel hopeful that their decision to pursue medicine will be personally rewarding (1b.6) and their salaries will stabilize (1a.1, 1a.5), but many low-income students experience doubt about whether they made the right career choice (1b.2, 1b.4, 1b.6), and feel stressed that they will be in debt for longer than they expected (1a.3, 1a.4, 1b.1, 1b.5). **Support low-income students to make values-based tradeoffs when considering a career in medicine.**

#### Design Opportunity

Support low-income students to make values-based tradeoffs when considering a career in medicine.

#### Insight 2

Medical schools lack the adaptive infrastructure to be welcoming to low-income students.

#### Description

Students face financial challenges from the moment they apply to medical school (theme 2a), a costly process that limits admissions options for low-income students due to their inability to pay for numerous application fees (2a.1) and expensive test preparation courses (2a.2, 2a.3). Once students begin medical school, they feel unsupported in their varied responsibilities towards their families (theme 2b) and additional financial needs (theme 2c), requiring them to make tradeoffs with their education and personal lives (2b.2, 2c.1).

#### Design Opportunity 2

Develop flexible systems that can recognize and accommodate students’ complex financial needs during medical school.

#### Insight 3

Students worry about the impact that their medical school debt has on their present and future families, which compounds feelings of guilt and anxiety.

#### Description

For students who need to take loans, the decision to pursue a career in medicine is a collective investment with their families. Students feel guilty about the sacrifices their families have to make for the sake of their career (theme 3a) and feel pressure to continue to provide financially for their family while having debt (theme 3b). Students are stressed about acquiring more debt throughout their training (3a.1) and the impact that has on loved ones who are dependent on them (3a.4, 3a.5, 3b.2), especially with respect to ensuring their financial security in the future (3b.4).

#### Design Opportunity 3

Create an environment that acknowledges and accounts for the burden of responsibility that students face towards their families.

#### Insight 4

Without the appropriate education about loans, the stress of debt is exponentially worse.

#### Description

Students feel the greatest fear around loans when they do not understand them, including the process of securing loans and paying off debt (theme 4a). Students are overwhelmed by their loan amounts (4a.5) and lack the knowledge or resources to manage their debt (4a.1, 4a.2), making them uncertain about how they will become debt-free in the future (4a.3, 4a.4). Students reported that various resources helped to alleviate those burgeoning fears (theme 4b), including financial aid counselors (4b.2, 4b.3) and physician role models (4b.5, 4b.6) that generally increase knowledge and skills related to debt management (4b.1).

#### Design Opportunity 4

Empower students to become experts in managing their debt by making loan-related resources more available and accessible.

#### Insight 5

The small, daily expenses are the most burdensome and cause the greatest amount of stress.

#### Description

Students with educational debt are mentally unprepared for the burden of managing their daily living expenses (theme 5a), causing them to make significant lifestyle adjustments in the hopes to ease their resulting anxiety (theme 5b). These costs are immediate and tangible, compared to tuition costs which are more distant and require less frequent management (5a.3) Students learn to temper their expectations for living beyond a bare minimum during medical school (5a.1, 5b.2, 5b.4) and develop strategies to ensure that their necessary expenses are as low as possible (5b.1, 5b.2, 5b.3, 5b.4).

#### Design Opportunity 5

Develop and distribute resources to support both short- and long-term financial costs for medical students.

#### Insight 6

Students view debt as a dark cloud that constrains their mental health and dictates their career trajectory.

#### Description

The constant burden of educational debt constrains students’ abilities to control their mental health (theme 6a) and pursue their desired career path in medicine (themes 6b & 6c). Students feel controlled by their debt (6a.3) and concerned that it will impact their [ability] to live a personally fulfilling life (6a.1, 6a.2, 6c.6), especially with respect to pursuing their desired medical specialties (6b.1, 6c.3, 6c.5, 6c.6). Students with scholarships, as opposed to loans, felt more able to choose specialties that prioritized their values rather than their finances (6c.1, 6c.2), an affordance that impacts long-term career growth and satisfaction.

#### Design Opportunity 6

Create a culture of confidence for managing debt and debt-stress among medical students.

## Discussion

This is the first multi-institutional national study to explore the impact of debt-related stress on medical students’ well-being in the United States. We used an innovative, mixed methods approach to better understand the factors that significantly affect debt-related stress, and propose opportunities for improving medical student well-being.

### URM Students

Analysis of survey results found that students who identify as URM are more likely to experience higher levels of debt-related stress than non-URM students. Our study also found that among all higher debt burden categories, debt levels were higher for URM students, findings consistent with studies that have shown the disproportionate burden of debt among URM students.^1^ Our semi-structured interviews illuminated that students from low-income backgrounds feel unsupported by their medical schools in these varied financial stressors that extend beyond tuition costs (insight 2), leaving their needs unmet and increasing financial stress over time: *“We don’t have different socio-economic classes in medicine because there’s constantly a cost that [isn’t] even factored into tuition cost [and] that we can’t take student loans for.”* Many URM students feel especially stressed by their financial obligations towards their families (insight 3), and describe the decision to enter into medicine as one that is collective (*“the family’s going to school”*) rather than individual, placing additional pressure on themselves to succeed in their career: “*Being of low SES, the most significant stressor for me is the financing of medical school and the pull of responsibility for my family.”* Several other studies from the literature confirm that students who identify as URM and first generation college or medical students are at higher risk for financial stress compared to their counterparts,^7^ and report that they feel as though it is their responsibility to honor their families through their educational and career pursuits.^23^ Our study demonstrates and describes how low-income and URM students face numerous financial barriers in medical school, resulting in medical trainees that are less diverse than the patient populations they are serving.^1,8^

### Debt Amount

Our quantitative analysis found that students with debt amounts over $100,000 are at much higher risk for experiencing severe stress than students with debt less than that amount. Although this finding may seem intuitive, it is important to highlight the degree to which this risk differs between these two cohorts. Students with debt amounts between $20,000 and $100,000 are approximately 5 times more likely to experience high stress than students with debt less than $20,000, while students with debt amounts over $100,000 are approximately 13 times more likely to experience severe stress when compared to the same cohort. Interview participants describe that the more debt they have, the less hopeful they feel towards achieving financial security (insight 1): *“There are other healthcare professionals that will not accrue the same amount of loans that we will, and then may or may not have the same salary or privileges […] makes me question, did I do the right thing?”* Students internalize this rising stress so as not to shift the feelings of guilt onto their families (insight 3), thereby compounding the psychological burden associated with large amounts of debt (insight 6): *“As long as you’re in debt, you’re owned by someone or something and the sooner you can get out of it, the better; the sooner I can get started with my life.”*

### Pre-Clinical Students

According to our survey analysis, students who are in their pre-clinical years are at higher risk for stress than students in their clinical years. Our interview findings from insight 4 suggest that students feel initially overwhelmed and unsure about what questions to ask (*“One of my fears is that I don’t know what I don’t know”)* or how to manage their loans so that it doesn’t have a permanent impact on their lives: *“The biggest worry is, what if [the debt] becomes so large that I am never able to pay it off and it ends up ruining me financially.”* Pre-clinical students may therefore feel unsure or ill-equipped to manage their loans, making them feel overwhelmed by the initial stimulus of debt. By the time students reach their clinical years, they may have had time to develop strategies for managing stress, acquire more financial knowledge, and/or normalize the idea of having debt.

### Medical School Characteristics

Our survey analysis found several risk factors related to medical school characteristics. First, we found that students who attended a private school were at higher risk for debt-related stress than students who attended a public school. Not only is the median 4-year cost of attendance in 2023 was almost $100,000 higher in private compared to public medical schools,^24^ but it is also the case that financial aid packages are more liberally available for public schools due to state government funding.^25^ This not only relieves students from having higher amounts of debt, but it also creates a more inclusive cohort of medical students. Insight 2 from our interviews reveal that private medical schools without the infrastructure that can adapt to students’ varying needs (e.g. financial aid vs loans).

Another characteristic that was found to be a risk factor for debt stress was attending a medical school on the West Coast (compared to a non-coastal school.) This was a surprising finding given that tuition rates for both private and public schools on the West Coast are no higher than those in other regions.^17^ The distribution of survey respondents did not vary significantly across regional categories, so no bias in sample size is suspected. A potential explanation might be that historically, students match for residency programs that are in their home state or not far from their home state.^26, 27^ Students may prefer to settle on the West Coast, and may be willing to take on more financial debt in pursuit of their long-term practice and lifestyle goals.

### Gap Year

Our quantitative analysis found that students who reported having considered taking a leave of absence for well-being purposes were at higher risk for debt-related stress. This cohort of students likely experience higher levels of stress as they are conscious of the negative impact it has on their life, and have already ruminated on leaving medical school. A study by Fallar et al. found that the period leading up to a leave of absence is particularly stressful for students because they are unfamiliar with the logistics of taking time off, and don’t feel as though leaving medical school is encouraged or normalized for students.^28^ An interview with a student who did a joint MD and PhD program expressed having more time for herself during her PhD program, and described using money for activities that could alleviate stress *(“I took figure skating during my PhD”)* rather than create more stress by compromising on their lifestyle during medical school (insight 5). More research may be needed to better understand and support students considering taking a leave of absence from medical school.

### Specialty Choice

Our study found that students with high debt stress pursue moderately competitive specialties compared to students with low debt stress. This may be explained by the fact that low debt stress gives students the freedom to pursue minimally competitive specialties, which may be more fulfilling to them but typically have lower salaries. Insight 6 further elaborates upon this finding that students with high debt stress deprioritize specialties for which they are passionate in favor of higher paying specialties that might alleviate their debt: *“I love working with kids…but being an outpatient pediatrician just wasn’t going to be enough to justify the* [private school] *price tag.”* Students with lower debt stress describe having the freedom to choose specialties that align with their values, regardless of anticipated salary: *“Scholarships give me the freedom to do [specialties] that maybe are a little bit less well-paying in medicine.”* Interestingly, certain studies examining the relationship between specialty choice and debt stress have found that high debt stress is associated with a higher likelihood of pursuing a more competitive, and presumably higher paying, specialty.^5^ More research investigating the relationship between debt stress and specialty choice could illuminate opportunities for increasing a sense of agency and overall satisfaction among students for their career choices.

### Resource Utilization

In our exploration of potential protective factors against the effects of debt-related stress, our survey analysis found that the two variables measured (high mental health resource utilization and meeting with a counselor) did not have any impact on reducing debt-related stress. This finding is inconsistent with the literature, which considers these activities to promote general well-being among students but has never been studied in the context of debt-related stress.^13–15^ A potential explanation is that the survey questions that assessed these activities were imperfect. For example, the question of meeting with a counselor was not a standalone question, but instead, was at the bottom of a list of other wellbeing activities; therefore, students may have been fatigued by the time they got to the bottom of the list and not selected it. Additionally, our definition of “high” mental health resource utilization may have been perceived as too strict (i.e.: 80-100%) and perhaps we would have seen effects at lower percentages of utilization (i.e.: 40-60%). Despite this finding, students describe in their interviews that having access to certain resources such as financial knowledge and physician role models can help to alleviate stress by helping them feel confident in managing their loans in the immediate and more distant future (insight 4): *“I’ve had explicit discussions with physicians who went to med school, had debt, paid it off […] the debt hasn’t hindered their life in any way. I think that just makes me feel a lot calmer.”* This finding aligns with previous studies that suggest that financial knowledge, such as knowledge about loans and a payoff plan, confers confidence in students’ financial management.^11,12^ These factors are also aligned with previous studies that suggest financial optimism, such as with a physician role model who successfully paid off loans, are associated with less financial stress.^10^

### Next Steps

Our quantitative analysis of risk factors helped us to identify which areas might significantly impact debt-related stress among medical students, while our qualitative analysis provided more in-depth insight into those risk factors for more human-centered intervention design. The HCD process not only provides additional context from the perspective of medical students, but also proposes distinct design opportunities upon which interventions may be designed and tested. The six design opportunities outlined in this paper span distinct areas of impact across the ecosystem, and support the co-design of solutions with students and key stakeholders. Next steps for this project are to build from the design opportunities to conduct structured brainstorming sessions with a diverse cross-section of stakeholders. In HCD brainstorms, design opportunities are framed as “How Might We?” (HMW) questions, which serve as brainstorming prompts and encourage a breadth of different ideas within a well-defined topic. For example, the design opportunity in insight 2 might be reframed as: “HMW develop the adaptive infrastructure necessary to recognize and accommodate students’ complex financial needs?” While existing solutions may be appropriate – such as providing affordable MCAT preparation or subsidized application fees – creative brainstorming can produce novel and innovative solutions that may not otherwise exist. Solution concepts are then selected for iterative prototyping and evaluation with stakeholders, and are continuously refined and modified until the prototype reaches desirability and feasibility targets.

### Limitations

Our study had some notable limitations. One potential limitation is that our data collection occurred between 2019-2021 for this publication in 2023. Additionally, as described in the original study,^3^ a limitation of the MSWS is the inability to determine a response rate of students due to the survey distribution by medical student liaisons from each medical school; under the reasonable assumption that the survey was distributed to every US allopathic medical student, the response rate was estimated to have been 8.7%.^3^ An additional limitation is the potential for response bias.^3^ A limitation of the qualitative interviews is the potential for response bias among the interviewees. Although we purposely sampled, the students who accepted the invitation to interview may have been students with extreme views, either very negative views of debt or very neutral views of debt. Additionally, the interviewees were not representative of all possible financial situations, given that most students were from private schools, which typically have higher tuition rates. Also, all students had debt amounts in the middle and high categories, with none in the low category. Finally, our model of risk factors for debt-related stress suggested the presence of negative confounding factors, which exerted effects on specific variables (i.e.: pre-clinical year, West Coast) for which univariate analysis found no significant associations but multivariate analysis did. We did not perform further analysis to identify which variables served as the negative confounding variables.

In conclusion, our mixed methods, cross-sectional study exploring debt-related stress and its impact on US medical students’ wellbeing and professional development revealed a set of risk factors and design opportunities for intervention. By using a combined quantitative and qualitative HCD approach, we were able to develop a broad, in-depth understanding of the challenges and opportunities facing medical students with education debt. With these efforts to support the well-being and academic success of students at higher risk of debt-related stress, medical education institutions can develop and nurture a more diverse medical field that can best support the needs of future patients.

## Data Availability

All data produced in the present study are available upon reasonable request to the authors

## Acknowledgements

We thank the members of The Better Lab, including Devika Patel and Marianna Salvatori, for their support. We appreciate Pamela Derish (UCSF) for assistance in manuscript editing and the UCSF Clinical and Translational Science Institute (CTSI) for assistance in statistical analysis.

This publication was supported by the National Center for Advancing Translational Sciences, National Institutes of Health, through UCSF-CTSI Grant Number UL1 TR001872. Its contents are solely the responsibility of the authors and do not necessarily represent the official views of the NIH.

## Supporting Information

**S1 File. Interview Guide S2 File. Codebook**

**S1 Table. Summary statistics of people who identified as having high debt stress (−2), vs. low debt stress (−1 or 0).**

