## Supplemental Table 1 for "Medical students in distress: a mixed methods approach to understanding the impact of debt on well-being"

**S1 Table. Summary statistics of people who identified as having high debt stress (-2), vs. low debt stress (-1 or 0).**

| | Level of Stress Associated with Medical School Debt | Overall | Low (-1 or 0) | High (-2) |
|---|---|---|---|---|
| N | | 2,771 | 2,176 | 595 |
| MS Year | Pre-Clinical | 1463 (52.8) | 1146 (52.7) | 317 (53.3) |
| | Clinical | 561 (20.2) | 437 (20.1) | 124 (20.8) |
| | Gap Year/Other | 134 ( 4.8) | 102 ( 4.7) | 32 (5.4) |
| | Post-Clinical | 613 (22.1) | 491 (22.6) | 122 (20.5) |
| Gender | Male | 919 (33.2) | 734 (33.8) | 185 (31.1) |
| | Non-Male | 1850 (66.8) | 1440 (66.2) | 410 (68.9) |
| Marital Status | Never Married | 2411 (87.1) | 1902 (87.5) | 509 (85.5) |
| | Divorced/Widowed | 19 (0.7) | 13 (0.6) | 6 (1.0) |
| | Married | 338 (12.2) | 258 (11.9) | 80 (13.4) |
| Disability | No | 2481 (91.0) | 1951 (91.3) | 530 (90.0) |
| | Yes | 245 ( 9.0) | 186 ( 8.7) | 59 (10.0) |
| URM | No | 2391 (88.6) | 1907 (89.6) | 484 (84.6) |
| | Yes | 309 (11.4) | 221 (10.4) | 88 (15.4) |
| Debt Burden | <$20K | 728 (27.9) | 691 (34.1) | 37 ( 6.4) |
| | $20-100K | 884 (33.9) | 699 (34.5) | 185 (31.8) |
| | >$100K | 997 (38.2) | 638 (31.5) | 359 (61.8) |
| Specialty Competitiveness | Low | 1309 (48.2) | 1039 (48.6) | 270 (46.6) |
| | Moderate | 985 (36.2) | 751 (35.1) | 234 (40.4) |
| | High | 424 (15.6) | 349 (16.3) | 75 (13.0) |
| Specialty Category | Surgical | 413 (15.2) | 326 (15.2) | 87 (15.0) |
| | Medical | 1414 (52.0) | 1122 (52.5) | 292 (50.4) |
| | Mixed (Surgical/Medical) | 891 (32.8) | 691 (32.3) | 200 (34.5) |
| Degree Type | MD | 2648 (95.8) | 2094 (96.6) | 554 (93.1) |
| | DO | 115 (4.2) | 74 (3.4) | 41 (6.9) |
| School Category | Private | 1409 (51.0) | 1089 (50.2) | 320 (53.8) |
| | Public | 1354 (49.0) | 1079 (49.8) | 275 (46.2) |
| Region | Northeast | 1019 (36.9) | 817 (37.7) | 202 (33.9) |
| | West Coast | 718 (26.0) | 547 (25.2) | 171 (28.7) |
| | Non-Coastal | 1026 (37.1) | 804 (37.1) | 222 (37.3) |

| | | | | |
|---|---|---|---|---|
| City Character | Non-Metropolitan | 1272 (46.1) | 990 (45.7) | 282 (47.6) |
| | Metropolitan | 1485 (53.9) | 1175 (54.3) | 310 (52.4) |
| School Average Tuition | <$40K | 365 (13.4) | 299 (14.0) | 66 (11.3) |
| | $40 - $60K | 1745 (64.1) | 1366 (63.8) | 379 (65.0) |
| | >$60K | 614 (22.5) | 476 (22.2) | 138 (23.7) |
| Leave of Absence for Wellbeing | Never Considered | 2177 (78.8) | 1747 (80.5) | 430 (72.4) |
| | Considered | 478 (17.3) | 344 (15.9) | 134 (22.6) |
| | Have Taken | 109 (3.9) | 79 (3.6) | 30 (5.1) |
