## Supplemental File 1 for "Medical students in distress: a mixed methods approach to understanding the impact of debt on well-being"

**S1 File. Interview Guide**

### Effects of Debt on MS Wellbeing
### Semi-Structured Interview Guide

**General:**

- Tell me a little about yourself - Where are you from? What got you interested in medicine? Did you consider any other careers?
- How has your medical school experience been so far?

**Your Journey:**

- Tell me about your journey with thinking about the costs of going into medicine.
    - When did you first start thinking about debt? Did physician salary influence your decision to go into medicine? Did medical school tuition influence where you chose to go to medical school? What led you to the decision to choose to take student loans (take on debt)?
- Tell me about your experience of debt in medical school.
    - What effects has debt had on you personally? How has your stress with debt changed during your time in medical school? How do you imagine it will change as you continue through medical school?
- Tell me about an experience when you were particularly stressed about your debt or financial situation?
- Tell me about a time when your stress about debt was eased?
- How does your debt influence your current choices? How does it affect how you look at the future?
- What services are provided by your school that are related to debt and financial stress? How helpful did you find these services?
- What are your sources of financial support?

**Unique Experience:**

- In what ways do you think your experience with debt in medical school has been different from your colleagues? How about colleagues you might have outside of medicine?
- What aren't medical schools considering when they think of debt?

**Institution's Role:**

- How much do you think medical school should cost?
- What do you think is a reasonable amount of student debt for a person?
- How should tuition fees be spent by a school? Where do you think your tuition money is being spent currently?
- What roles do you think medical schools should have with respect to student debt?

**Effect of COVID:**

- How has COVID affected your experience with debt?
- How has COVID affected your costs?

**Solutions:**

- What do you wish was being done for you with respect to your debt that currently isn't?

- If you had all the resources in the world, how would you best tackle debt in medical school and the stress it causes?
    - What if you could only spend $100 per student, how would you best tackle it then? How would you change the system?

**Wrap Up:**

- How has it felt talking about this subject?
- Are there any questions that I should have asked you that I didn't?
