## Supplemental File 2 for "Medical students in distress: a mixed methods approach to understanding the impact of debt on well-being"

**S2 File. Codebook**

### Medical Student Debt, Interview Code Book

***Arranged by domains and codes (#'s)**

#### Journey to Being a Doctor

1. **Personal Journey into Medical School**
   - Significant events occurring prior to medical school that influenced or impacted student.
   - Examples: *international student came to American on her own where she was introduced to medicine as a possibility for her future, premature baby that had a lot of exposure to medical community throughout life and inspired her to become physician, other career options prior to medical school.*

2. **Career Expectations**
   - Thoughts or expectations of what a career in medicine would be like prior to entering field.
   - Examples*: financial expectations, workload, details of how time would be spent at work.*

3. **Financial Journey into Medical School**
   - Significant events occurring prior to medical school where costs, financials or debt considerations were at the focus.
   - Examples: *consideration of what medical schools to apply to base on costs, undergraduate debt, when costs of medical school were first considered.*

4. **Future Specialty Considerations**
   - Student's specialty plans after medical school and which factors influenced their decision; student's thoughts on specialties as it relates to finances/debt/etc.
   - Examples: *choosing or avoiding specialty due to the payouts of each, stress or tension between doing specialty that student really loves and worrying about payout.*

5. **Post-Medical School Payback Plan / Repayment Option Impressions:**
   - Student's plan for managing their debt after medical school is complete; students impression of pros and cons for repayment options available.
   - Examples: *deferment, debt forgiveness programs (military, income-based, underserved area), strategies for payments during residency.*

6. **Medical School During COVID**
   - Impressions of how COVID has impacted medical school (logistics, quality of education, value of education, impression of costs / financials during this time)
   - Examples: *petitions from students for school to lower cost of education, resentment about remote learning during pandemic, logistic considerations such as residency interviews over Zoom.*

#### Getting Through Medical School Finances

7. **Financial Aid Office**
   - Impression or anecdotes about student's interaction with their medical schools' financial aid office.
   - Examples: *communications with office, helpfulness for navigating the financial path in medical school*

8. **Loans / Scholarships / Money from Institution**
   - Logistics of applying for institutional or federal loans; availability and distribution of money; loan structures and terms

- Examples: *FAFSA paperwork, loan structures for limits, loan interest structure, deferment options, availability of funds*

9. **Paying Your Way**
   - How student navigates planned or unplanned expenses during medical school
   - Examples: *Borrowing money from parents, dependence on spouse, using loans, expense workarounds.*

10. **Financial Support System**
   - Individuals that student relies on for financial support; Lack of individuals available to student for financial support; Methods of support that are offered outside of providing direct payment.
   - Examples: *partner research best plan for loan repayment option, family helping to pay for expenses during school, ways in which student can't turn to others for financial assistance.*

#### Institution's Role

11. **Impression of Institution**
   - How student views the motives, workings and considerations of their medical school; student expectations of their schools' costs, commitments and obligation to them.
   - Examples: *motives of administration / school, impression of endowments and availability of money for students, expectations that school should provide funds for students*

12. **Allocation of Tuition Money**
   - Student's impressions of how money is spent at their medical school; thoughts on how money would better be spent.
   - Examples: *ambiguity of money allocation, distrust that tuition money doesn't go to pay for student's actual education, desires on how funds should better be allocated.*

13. **Impression of Healthcare / Medical Education**
   - Student's impression of how the healthcare system or medical education system at large influences the cost of medical school.
   - Examples: *inflated cost of physician salary, impressions of who the stakeholders are that influence medical school cost.*

14. **Needed Resources at School / Solutions**
   - Wants / Solutions directly stated or implied by students to help alleviate the impact of debt on medical student wellbeing.
   - Examples: *loan restructuring, changing the cost of medical school, better programs to prepare students for how to tackle debt incurred.*

#### Emotional Toll of Debt

15. **Sources of Financial Stress / Fears**
   - Specific financial situations or circumstances that increase student's stress; student fears involving costs, debt, or the impact each of these has or will have on their lives.
   - Examples: *needing to provide for family, fears about uncertainty about the future, emergency situations that require unaccounted for funds.*

16. **Debt Stress Manifestations**
   - Description of what debt stress looks like in a student's life; emotional response to debt.
   - Examples: *loss of sleep, anxiety, resentment of the school, exhaustion from thinking about finances constantly, increased pressure to produce / succeed.*

**17. Coming from Low Socioeconomic Background**

- Examples of the unique journey a student from a low socioeconomic background contends with.
- Example: *unforeseen costs, toll of expenses, lack of safety network, sense of community or isolation.*

**18. Coping Mechanisms:**

- Ways in which medical students cope with the stress of finances during medical school; helpful resources for student to deal with stress of finances.
- Examples: *avoidance, yoga, sharing with community, proactive efforts to understand / deal with debt.*
